# The Australian Eye and Ear Health Survey (AEEHS): study protocol for a population-based cross-sectional study

**DOI:** 10.1101/2023.05.25.23290454

**Authors:** Richard Kha, Oonagh Macken, Paul Mitchell, Gerald Liew, Lisa Keay, Colina Waddell, Eleanor Yang, Vu Do, Tim Fricke, John Newall, Bamini Gopinath

## Abstract

**Introduction:** Vision and hearing impairments are highly prevalent and have a significant impact on physical, psychological and social wellbeing. There is a need for accurate, contemporary national data on the prevalence, risk factors and impacts of vision and hearing loss in Australian adults.

**Objectives:** The Australian Eye and Ear Health Survey (AEEHS) aims to determine the prevalence, risk factors and impacts of vision and hearing loss in both Aboriginal and Torres Strait Islander and non-Indigenous older adults.

**Methods and analysis:** The AEEHS is a population-based cross-sectional survey which will include 5,000 participants (3250 non-Indigenous aged 50 years or older and 1750 Aboriginal and Torres Strait Islander people aged 40 years or older) from 30 sites covering urban and rural/regional geographic areas, selected using a multi-stage, random cluster sampling strategy. Questionnaires will be administered to collect data on socio-demographic, medical, ocular and otological history. The testing battery includes assessment of blood pressure, blood sugar, anthropometry, visual acuity (presenting, unaided, pinhole and best-corrected), refraction, tonometry, slit lamp and dilated eye examination, ocular imaging including optical coherence tomography (OCT), OCT-angiography and retinal photography, and automated visual fields. Audiometry, tympanometry and video otoscopy will also be performed. The primary outcomes are age-standardised prevalence of cause-specific vision and hearing impairment. Secondary outcomes are prevalence of non-blinding eye diseases (including dry eye disease), patterns in health service utilisation, universal health coverage metrics, risk factors for vision and hearing impairment, and impact on quality of life.

**Ethics:** The protocol for this study was approved by the University of Sydney Human Research Ethics Committee (HREC-2020/818) and Australian Institute of Aboriginal and Torres Strait Islander Studies (AIATSIS) Research Ethics Committee (HREC-EO303-20211008).

**Dissemination of results:** Our findings will be disseminated through presentation at meetings and peer-reviewed publications. Findings will also be widely disseminated by project partners with the aim of improving public health policy directives and equitable service delivery to prevent avoidable vision and hearing impairment in Australia.

## Introduction

Vision and hearing impairment have been found to be highly prevalent chronic conditions that have significant impacts on health and wellbeing.^1^ As such, contemporary studies to establish the prevalence, risk factors and impact of vision and hearing loss among Aboriginal and Torres Strait Islander people and non-Indigenous Australians are needed. Globally, 285 million individuals are visually impaired, of whom 39 million are blind.^2^ In Australia, there are 840,000 individuals estimated to be living with vision loss, and this number is expected to exceed 1.04 million by 2030.^3^ Furthermore, there are currently 3.6 million Australians estimated to be living with hearing loss and this is projected to rise to 7.8 million by 2060.^4^

The social, economic and health system costs of vision and hearing impairment are significant. In Australia, the economic impact of vision and hearing loss is $27.6 and $11.8 billion per year, respectively.^3, 5, 6^ Vision and hearing loss are strongly associated with reduced quality of life, mental and physical health, as well as increased mortality and social isolation.^6–8^ Over 90% of vision and hearing loss can be prevented or treated through identification of risk factors, early diagnosis and management.^9, 10^ Hence, accurate age– and sex-specific data for visual and hearing impairment over time are critical.

In addition to the need for national prevalence data in Australia, there is also a need to identify population subgroups with greater burden of vision and hearing loss. Visual and hearing impairment rates have previously been estimated to be three and ten times higher among Aboriginal and Torres Strait Islander people when compared to non-Indigenous communities, respectively.^11–13^ Furthermore, global studies have found that eye and ear conditions are more prevalent in people living in rural compared to urban geographical areas.^14, 15^ As such, there is a need for contemporary data in Australia to determine the extent and sources of disparities in age-specific risk of visual and hearing impairment between these populations.

This study will be the second nationwide population-based survey on eye health and first nationwide population-based survey on ear health in Australia. The results of the AEEHS will provide valuable information regarding Australia’s current eye and ear health status, outlining gaps in treatment, diagnosis and prevention, and assessing potential links between eye disease, hearing loss and critical health and/or social outcomes. This will assist in developing public health policy initiatives, guiding future resource allocation and delivery of vision and hearing services to better prevent and manage vision and hearing loss. Furthermore, it will be possible to track changes in eye health since the first National Eye Health Survey (NEHS1) in 2016 to assess whether progress has been made to lessen the burden of vision loss.

## Specific objectives

– To establish the current prevalence, risk factors and impacts of visual and hearing impairment in Australian adults.
– To identify the modifiable and non-modifiable risk factors for vision and hearing loss in adults.
– To analyse trends in eye health since the 2016 NEHS1 and report key metrics for universal eye health coverage.
– Compare the prevalence of visual and hearing loss between Aboriginal and Torres Strait Islander people and non-Indigenous Australians, and between each urbanisation level.

## Methods

The Australian Eye and Ear Health Survey is a cross-sectional study which aims to recruit and examine a total of 5,000 participants across 30 randomly selected sites in Australia. Sample size calculation assumed a similar prevalence of visual impairment in the NEHS1.

The protocol for this study was approved by the University of Sydney Human Research Ethics Committee (HREC-2020/818) and Australian Institute of Aboriginal and Torres Strait Islander Studies (AIATSIS) Research Ethics Committee (HREC-EO303-20211008).

## Sample Size

A total of 5,000 participants will be surveyed across 30 randomly selected sites in Australia. The target population includes 1,750 Indigenous Australians (aged 40 years and older) and 3,250 non-Indigenous Australians (aged 50 years and older). The proportion of Indigenous populations is oversampled 10-fold, as Aboriginal and Torres Strait Islander people represent 3.2% of the general Australian population.^16^

A sample size of 5000 will provide 99% power to detect 2.87% differences in visual impairment prevalence in Indigenous Australians and 90% power in non-Indigenous Australians. There will also be over 95% power to detect a linear trend of decreasing vision impairment in the non-Indigenous cohort (using the Cochran-Armitage test for trend in proportions with continuity correction). These figures assume a statistical significance level of 0.05 and are based on the reported prevalence among Indigenous and non-Indigenous participants in the first National Eye Health Survey (NEHS1) respectively. The same sample size will provide over 90% power to accurately obtain hearing impairment prevalence.

## Site Selection

Multi-stage, random cluster sampling will be used to select 30 sites to provide a sample of 3250 non-Indigenous Australians aged 50 years and older and 1750 Aboriginal and Torres Strait Islander Australians aged 40 years or older. 30 recruitment sites were required based on this sample size, and an expected cluster of 100 non-Indigenous individuals and 50 Indigenous individuals in each site. Aboriginal and Torres Strait Islander participants have a younger inclusion age criteria because major eye and hearing conditions such as diabetic retinopathy and glue ear have an earlier onset and faster rate of progression in these communities.

The site sampling for the AEEHS was based on Statistical Area Level 2 (SA2) regions, which are defined in the Australian Statistical Geography Standard (ASGS).^17^ Statistical Areas Level 2 (SA2) are medium-sized general-purpose areas built up from multiple whole Statistical Areas Level 1 (SA1). A total of thirty SA2 sites were stratified by state and territory to ensure that the sample was reflective of the geographic distribution of both Indigenous and non-Indigenous older Australians (Table 1).

**Table 1.**
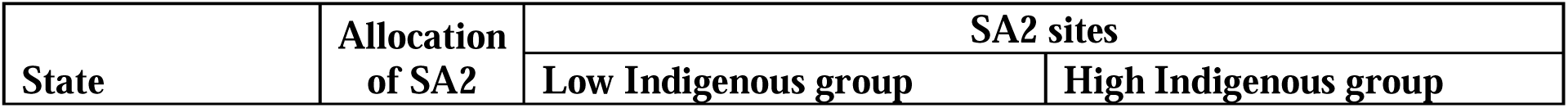
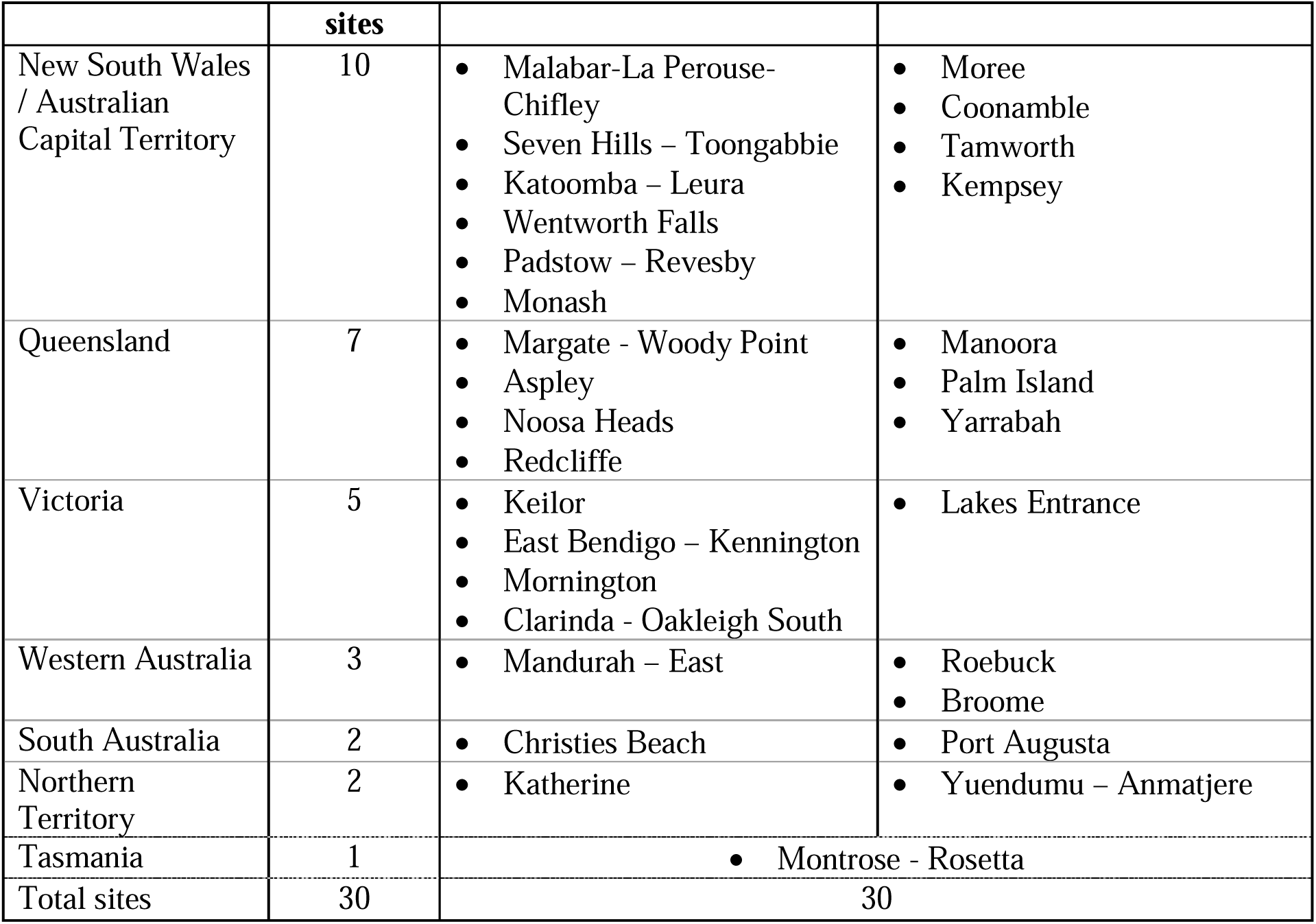
Proposed examination sites for the AEEHS

A second level of stratification was employed at the state/territory level based on Indigenous status. SA2 sites that had 80% or higher Indigenous population were categorised in the ‘high Indigenous’ proportion group whilst the remainder was allocated to the ‘low Indigenous’ proportion group’. An initial level of stratification was based on remoteness area defined by the AGSG^17^ and based on the distribution of the Australian population, 19 Major City, seven Inner Regional, three Outer Regional and one Remote/Very Remote sites were sought. Within each selected SA2 region, two to three SA1 regions were selected as the recruitment sites.

### Inclusion criteria

⍰ Non-Indigenous Australians aged ≥50 years, or Indigenous Australians aged ≥40 years;
⍰ Australian citizen or permanent resident
⍰ Able to provide written informed consent
⍰ Residing within selected SA2 boundaries

## Recruitment

Recruiters will go door-to-door at each site to recruit participants in each randomly selected SA1. An information pack containing a letter and information pamphlet outlining the study and a statement that recruiters will doorknock at their residence, will be left in each mailbox. Recruiters use a standardised script and screen for eligibility based on the inclusion criteria. Eligible residents will be invited to participate and those who agree will be given an appointment card with the date, time and venue. Socio-demographic information including their age, gender, date of birth, address, contact details and Indigenous status will be recorded on the online database REDCap. Each home will be approached two times and any individuals who have declined to participate will not be recontacted. Residents who were not present following both door-knock attempts will be deemed non-contactable. Additional modes of recruitment will be implemented as appropriate through discussion with community leaders in order to adhere to cultural norms. These may include word of mouth, media announcements, and community engagement.

## Data collection and testing protocol

Participant examinations will take place onsite and consists of four stations. Consenting participants will undergo general health and eye assessment at Stations 1-3, followed by an ear health assessment at Station 4. The examination protocol is summarised in Figure 1. Verbal and written feedback in the form of a report is provided to each participant with appropriate referrals to an optometrist, audiologist and/or local GP if abnormalities are present. The testing protocol will take approximately 60-90 minutes per patient. A voluntary take-home questionnaire will be provided to all participants to complete in their own time. This questionnaire gathers additional information about the participant’s self-reported vision and hearing, health, mental wellbeing and lifestyle. It includes:

– National Eye Institute: Visual Function Questionnaire^18^
– Ocular Surface Disease Index^19^
– Health Outcomes (EuroQol Group EQ-5D-5L, EQ-5D Visual Analogue Scale)^20^
– Fatigue Severity Scale^21^
– Incidental and Planned Exercise Questionnaire^22^
– Food Frequency Questionnaire
– Questions about exposure to specific illnesses, noise and ototoxic chemicals, as well as history of ear infections

**Figure 1.**
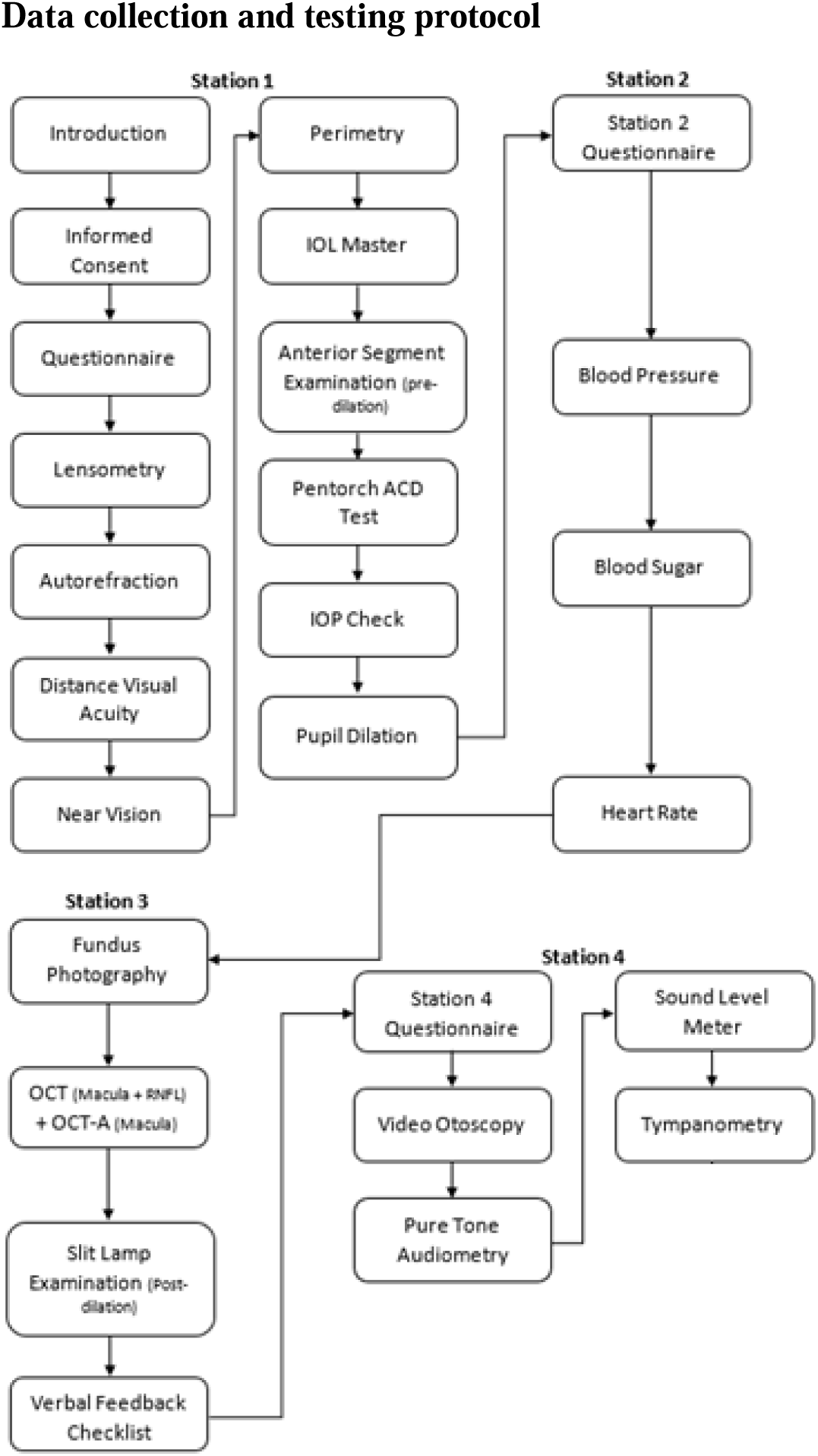
Flow chart of the AEEHS examination protocol

**Figure 2.**
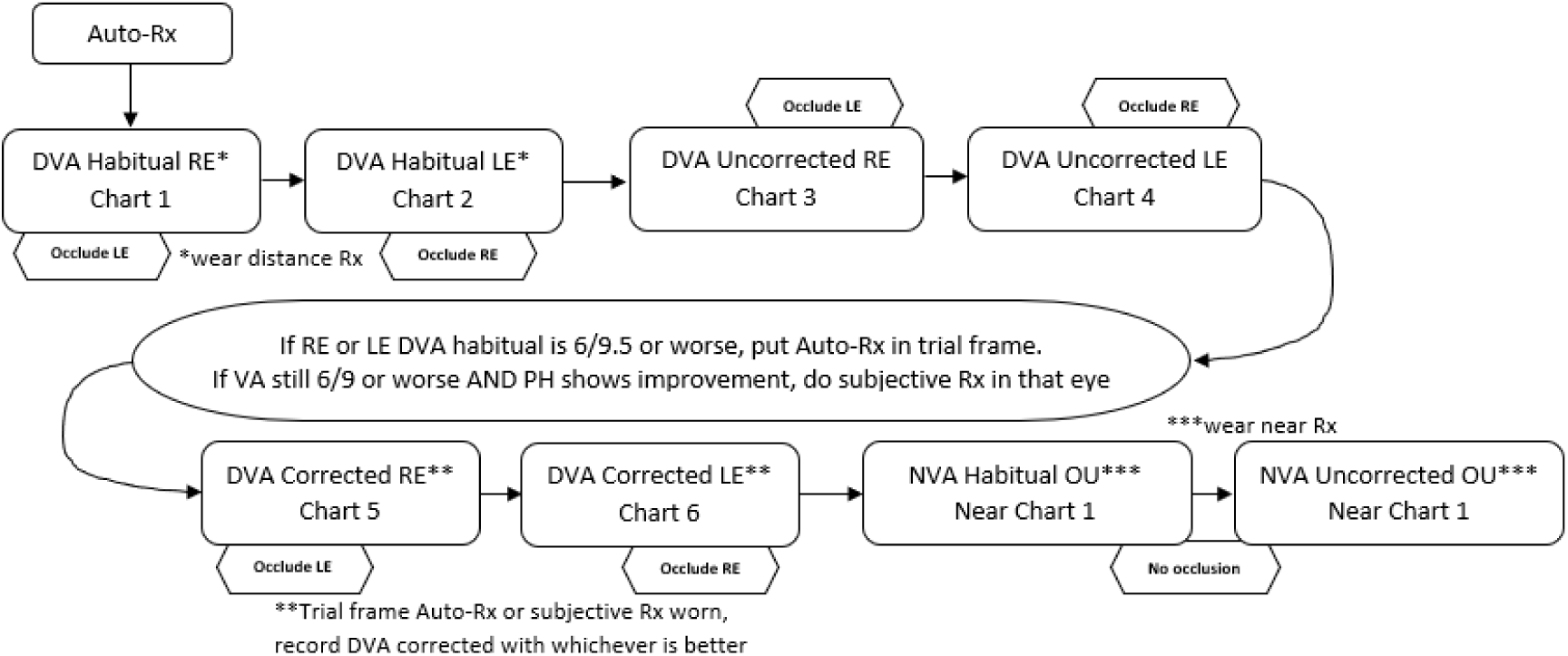
Flow chart for assessment of vision and refraction

### Station 1

⍰ Distance visual acuity (DVA) – Presenting DVA will be measured monocularly using the ETDRS LogMAR chart in well-lit room conditions at a distance of 2m with spectacles or contact lenses, if worn. If VA is less than 6/60, a Snellen chart will be used to test optotypes from 5/60 to 1/60. If no letters can be identified on the chart, VA will be assessed as counting fingers, hand movements, perception or no perception of light. Unaided DVA will also be assessed, followed by any improvement with pinhole. Vision impairment is defined as VA ≤ 6/12 whereas blindness is defined VA ≤ 6/60.
⍰ Refraction – If habitual DVA ≤ 6/9.5 and improves with pinhole, DVA will be measured using trial frames with autorefraction readings as the starting point, followed by subjective refinement of sphere, cylinder and axis until best corrected visual acuity is obtained.
⍰ Near vision acuity (NVA) – Habitual NVA is tested binocularly using a logMAR near vision chart at 40cm with spectacles, if worn. Unaided NVA will also be assessed.
⍰ Lensometry – the power of participants’ spectacles will be measured using the Zeiss VISULENS 550. This provides information on refractive error coverage.
⍰ Ocular biometry – non-contact partial coherence laser interferometry (Zeiss IOLMaster) will be used to measure axial length, anterior chamber depth, corneal curvature, lens and central corneal thickness.
⍰ Autorefraction – Zeiss VisuREF 150 autorefractor will be used to objectively measure spherical and cylindrical refractive error.
⍰ Pre-dilation anterior segment examination – gross assessment of iris colour, pterygium, pupil, eyelid abnormalities and anterior chamber depth will be performed using a bright LED pentorch.
⍰ Tonometry – intraocular pressure (IOP) will be measured using the iCare tonometer. If IOP>25mmHg or difference between eyes ≥5mmHg, a repeat reading will be taken using Goldmann applanation tonometry (Haag-Streit).
⍰ Visual fields – a 24-2 SITA Faster visual field examination of each eye will be performed using the Zeiss Humphrey Field Analyser III. This is used to determine visual field abnormalities which can reflect glaucoma, neurological disease and other retinal conditions.
⍰ Pupil dilation – tropicamide 1.0% will be instilled in each eye at the end of Station 1 to dilate pupils of consenting patients to optimise eye health assessment and imaging.

### Station 2

⍰ Questionnaire – includes questions on demographics including educational/occupational status, income, country of birth, smoking status, use of medications, and medical, surgical, ocular history (e.g. history of cataract, AMD, glaucoma, diabetic retinopathy and previous ocular surgeries). These questions are important for linking risk factors with eye disease.
⍰ Anthropometry – Height, weight and waist circumference will be measured using a height stand, Wedderburn electronic scale and measuring tape, respectively.
⍰ Blood pressure and heart rate will be measured using the Omron HEM 700.
⍰ Blood glucose will be measured using a finger prick test using the Accu-Chek glucose meter.
⍰ Cognitive function will be measured using the Montreal cognitive assessment (MoCA) for subset of participants over 65 years.

### Station 3

⍰ Optical coherence tomography (OCT) – This will be the first nationwide survey to use OCT imaging technology. The Zeiss Cirrus 6000 will be used to examine the optic nerve head, macula and retinal nerve fibre layer in both eyes. OCT is widely used for the diagnosis of retinal conditions including AMD, DR, macular holes and optic nerve disorders including glaucoma. Wide-field OCT-angiography scans will also be taken to non-invasively detect principal biomarkers that underlie neovascular age-related macular degeneration and diabetic retinopathy. This will be the first population survey worldwide to use OCT-angiography on a nationally representative sample. Anterior segment imaging will be used to assess anterior chamber angle on participants with Pentorch Anterior Chamber Depth Grade 1.
⍰ Fundus photography – Ultra-widefield colour (200^°^ field of view) and fundus autofluorescence retinal images of each eye will be captured using the Zeiss Clarus 700 retinal camera. An external image of each eye will also be taken using the camera to observe the lens through the dilated pupil. Photographs will then be graded using BMES and Wisconsin grading protocols by masked, trained ophthalmologists.
⍰ Post-dilation slit lamp examination – the slit lamp (Haag-Streit BM900) will be used to examine the anterior segment for signs of trachoma trichiasis, pseudoexfoliation, pigment dispersion, corneal opacities and cataract. The Lens Opacities Classification System III (LOCS III) will be used to grade the type and severity of cataract. It consists of a series of slit-lamp images for grading nuclear, cortical and posterior subcapsular cataract. Fluorescein dye is instilled last to assess tear break-up time and corneal staining using the Oxford staining score in order to detect dry eye disease.

### Station 4

⍰ Questionnaire – includes an interviewer-administered assessment on otological history, access to services for their hearing loss, and use of hearing devices.
⍰ Hearing Handicap Inventory for the Elderly – questionnaire assessing self-perceived hearing handicap in participants with pre-existing hearing loss.
⍰ International Outcome Inventory for Hearing Aids – questionnaire evaluating outcomes of aural rehabilitation in participants who utilise amplification devices
⍰ Video Otoscopy – visual inspection of the ear canal and tympanic membrane will be performed using the MedRx video otoscope. Observations are categorised as normal or abnormal tympanic membrane and ear canal presentations. Digital photographs will be taken for each ear and graded by masked hearing professionals.
⍰ Pure-tone audiometry – will be conducted using the Advant A2D Audiometer with passive noise reducing headphones (DD65v2) using a Hughson-Westlake staircase procedure. Noise levels will be monitored using a Burel and Kjaer type 2250 sound level meter.
⍰ Tympanometry – will be undertaken using the Amplivox Otowave 102-1. Results will record numerical data including static compliance, gradient, tympanometric peak pressure and ear canal volume (ECV). Responses will be typified using the Jerger classification system for tympanograms.

## Data storage

The data collected for analysis will be stored via University of Sydney REDCap, a secure, web-based application for managing online databases. Imaging data will be stored on a secure online server at Westmead Institute of Medical Research.

## Data analysis

95% CIs for age-standardised prevalence of diseases will be calculated using the normal approximation with Australian population using 2021 Census data. ANOVA will be used to compare the mean among groups of normally distributed parameters. Chi-square tests will be used to compare proportions. Multivariate logistic regression models will be used to examine the association of potential risk factors with disease. A two-tailed p-value <0.05 will be considered statistically significant.

## Dissemination of findings

A one-page summary of the study findings will be shared with the participants via their choice of post or email. Study completion is expected by the end of 2024 with data analysis and reports following in the first half of 2025. Findings will be disseminated through publications in peer-reviewed journals and presentations at conferences. Findings will also be widely disseminated by project partners with the aim of improving public health policy directives and equitable service delivery to prevent avoidable vision and hearing loss in Australia.

## Data Availability

All data produced in the present study are available upon reasonable request to the authors

## References

1. Fisher D, Li CM, Chiu MS, Themann CL, Petersen H, Jónasson F, et al. Impairments in hearing and vision impact on mortality in older people: the AGES-Reykjavik Study. Age Ageing. 2014;43(1):69–76.

2. Trends in prevalence of blindness and distance and near vision impairment over 30 years: an analysis for the Global Burden of Disease Study. Lancet Glob Health. 2021;9(2):e130–e43.

3. Vision 2020 Australia. Vision 2020 Australia Federal Budget Submission 2022-23 [Internet]. Melbourne: Vision 2020 Australia; 2022 [cited 2022 Jan 22]. Available from: https://www.vision2020australia.org.au/wp-content/uploads/2022/02/Vision-2020-Australia-Federal-Budget-Submission-22-23.pdf.

4. Taylor S, Cairns A, Solomon S, Glass B. Community pharmacist interventions in ear health: a scoping review. Prim Health Care Res Dev. 2021;22:e63.

5. Begg S, Vos T, Barker B, Stevenson C, Stanley L, Lopez AD. The burden of disease and injury in Australia 2003 [Internet]. Canberra: AIHW; 2007 [cited 2022 Jan 22]. Available from: https://www.aihw.gov.au/reports/burden-of-disease/burden-of-disease-injury-australia-2003/summary.

6. Davis A, McMahon CM, Pichora-Fuller KM, Russ S, Lin F, Olusanya BO, et al. Aging and Hearing Health: The Life-course Approach. Gerontologist. 2016;56 Suppl 2(Suppl 2):S256–67.

7. Assi L, Chamseddine F, Ibrahim P, Sabbagh H, Rosman L, Congdon N, et al. A Global Assessment of Eye Health and Quality of Life: A Systematic Review of Systematic Reviews. JAMA Ophthalmology. 2021;139(5):526–41.

8. Ehrlich JR, Ramke J, Macleod D, Burn H, Lee CN, Zhang JH, et al. Association between vision impairment and mortality: a systematic review and meta-analysis. Lancet Glob Health. 2021;9(4):e418–e30.

9. Burton MJ, Ramke J, Marques AP, Bourne RRA, Congdon N, Jones I, et al. The Lancet Global Health Commission on Global Eye Health: vision beyond 2020. Lancet Glob Health. 2021;9(4):e489–e551.

10. Olusanya BO, Davis AC, Hoffman HJ. Hearing loss: rising prevalence and impact. Bull World Health Organ. 2019;97(10):646-a.

11. Yashadhana A, Fields T, Blitner G, Stanley R, Zwi AB. Trust, culture and communication: determinants of eye health and care among Indigenous people with diabetes in Australia. BMJ Glob Health. 2020;5(1):e001999.

12. Foreman J, Xie J, Keel S, van Wijngaarden P, Sandhu SS, Ang GS, et al. The Prevalence and Causes of Vision Loss in Indigenous and Non-Indigenous Australians: The National Eye Health Survey. Ophthalmology. 2017;124(12):1743–52.

13. Access Economics. Listen Hear! The economic impact and cost of hearing loss in Australia [Internet]. Sydney: Access Economics; 2006 [cited 2022 Jan 22]. Available from: https://hearnet.org.au/wp-content/uploads/2015/10/ListenHearFinal.pdf.

14. Li Y, Hu QR, Li XX, Hu YH, Wang B, Qin XY, et al. Visual acuity of urban and rural adults in a coastal province of southern China: the Fujian Eye Study. Int J Ophthalmol. 2022;15(7):1157–64.

15. Swanepoel D, Clark JL. Hearing healthcare in remote or resource-constrained environments. J Laryngol Otol. 2019;133(1):11–7.

16. Australian Institute of Health and Welfare. Profile of Indigenous Australians [Internet]. Canberra: Australian Institute of Health and Welfare, 2022 [cited 2022 Jan 22]. Available from: https://www.aihw.gov.au/reports/australias-health/profile-of-indigenous-australians.

17. Australian Bureau of Statistics. Australian Statistical Geography Standard (ASGS) Edition 3 [Internet]. Canberra: Australian Bureau of Statistics; 2021 [cited 2023 Jan 26]. Available from: https://www.abs.gov.au/statistics/standards/australian-statistical-geography-standard-asgs-edition-3/latest-release.

18. Mangione CM, Lee PP, Pitts J, Gutierrez P, Berry S, Hays RD. Psychometric properties of the National Eye Institute Visual Function Questionnaire (NEI-VFQ). NEI-VFQ Field Test Investigators. Arch Ophthalmol. 1998;116(11):1496–504.

19. Schiffman RM, Christianson MD, Jacobsen G, Hirsch JD, Reis BL. Reliability and validity of the Ocular Surface Disease Index. Arch Ophthalmol. 2000;118(5):615–21.

20. Hinz A, Kohlmann T, Stöbel-Richter Y, Zenger M, Brähler E. The quality of life questionnaire EQ-5D-5L: psychometric properties and normative values for the general German population. Qual Life Res. 2014;23(2):443–7.

21. Rosti-Otajärvi E, Hämäläinen P, Wiksten A, Hakkarainen T, Ruutiainen J. Validity and reliability of the Fatigue Severity Scale in Finnish multiple sclerosis patients. Brain Behav. 2017;7(7):e00743.

22. Delbaere K, Hauer K, Lord SR. Evaluation of the incidental and planned activity questionnaire (IPEQ) for older people. Br J Sports Med. 2010;44(14):1029–34.

